# Prevalence of protective hepatitis B surface antibody levels and associated factors among women admitted for delivery at Kawempe National Referral Hospital, Uganda: A cross-sectional study

**DOI:** 10.64898/2026.11.10.26367773

**Authors:** Emilly Kembabazi, Moses Ocan, Hood Ibanda

## Abstract

Mother-to-child transmission sustains the burden of hepatitis B virus (HBV) infection in sub-Saharan Africa, yet little is known about whether women giving birth in Uganda are protected. We estimated the prevalence of protective hepatitis B surface antibody (anti-HBs) levels, described serological profiles and identified associated factors among women admitted for delivery at Kawempe National Referral Hospital, Kampala. In this cross-sectional study, 418 women were selected by systematic random sampling in 2024 and interviewed about socio-demographic characteristics, hepatitis B testing, vaccination and exposures. Anti-HBs was quantified by electrochemiluminescence immunoassay (protective: ≥10 mIU/mL); other HBV markers were screened with a rapid test. Prevalence ratios were estimated with modified Poisson regression. Overall, 264 women (63.2%) reported vaccination: 203 during the current pregnancy and 61 before it only. Protective levels were found in 322 women (77.0%; 95% confidence interval [CI] 72.8–80.8): 50.6% of never-vaccinated women, 82.0% of those vaccinated before the current pregnancy only and 95.6% of those vaccinated during it. After adjustment, vaccination before the current pregnancy only (adjusted prevalence ratio 1.54; 95% CI 1.24–1.89) and during it (1.86; 95% CI 1.57–2.20) were associated with protection; no socio-demographic or behavioural factor was. Eighty-two women (19.6%) had no serological evidence of immunity or infection; 65 had never been vaccinated. Of 300 women who reported a previous negative test, 50 (16.7%) had never been vaccinated, and 68 of 250 vaccinated women with known completion status (27.2%) had not completed three doses, mostly because they were not told to return or forgot. The rapid test detected hepatitis B surface antigen in 21 women (5.0%), but discordant marker patterns suggest some false positives. One in five women reached delivery susceptible to HBV, largely because a negative test was not followed by vaccination. Linking every negative antenatal test to vaccination could close much of this gap.

## Introduction

Hepatitis B virus (HBV) infection remains a leading cause of cirrhosis and liver cancer worldwide. In 2022, an estimated 254 million people were living with chronic HBV infection and 1.1 million died of HBV-related causes [1]. Global prevalence of hepatitis B surface antigen (HBsAg) was estimated at 3.2%, and only 46% of newborns received a timely birth dose of hepatitis B vaccine [2]. In highly endemic settings, most chronic infections are acquired at birth or in early childhood. Without prophylaxis, up to 90% of infants born to mothers who are positive for hepatitis B e antigen (HBeAg) acquire the infection, and around 90% of infants infected perinatally develop chronic hepatitis B [3,4]. The World Health Organization (WHO) therefore recommends a birth dose of hepatitis B vaccine within 24 hours of delivery, antenatal HBsAg testing and antiviral prophylaxis for eligible pregnant women [5,6].

A recent meta-analysis estimated the pooled HBsAg prevalence among pregnant women in Africa at 6.8%, and at 4.9% in East Africa [7]. In Uganda, a national serosurvey reported an HBsAg prevalence of 10.3% [8], and the 2016–2017 Uganda Population-based HIV Impact Assessment reported 4.1% among adults aged 15–64 years, with wide regional variation [9]. Among pregnant women, HBsAg prevalence has ranged from 11.8% in northern Uganda [10] to 2.1% in Lwengo District [11] and 1.2% in Mbarara Municipality [12]. Ugandan infants have received hepatitis B vaccine as part of the pentavalent vaccine from six weeks of age [10], and a national birth dose was introduced only in September 2025 [13]. Since 2015, Uganda has offered free hepatitis B screening for adults, with vaccination for those who test negative and treatment for those who are infected [14]. However, uptake and completion of the three-dose series are constrained by vaccine stock-outs, limited follow-up and vaccine hesitancy, including among pregnant women in Kampala [15,16].

A hepatitis B surface antibody (anti-HBs) concentration of at least 10 mIU/mL is the accepted correlate of protection [5,17]. Antibody response to vaccination varies with age, sex, body mass index, smoking, comorbidity and the number of doses received [18], and antibody concentrations decline over time after vaccination [19]. Among vaccinated healthcare workers in northern Uganda, 83% had protective anti-HBs levels [20], but data for pregnant women are scarce; in Mbarara, only 14% of pregnant women had serological evidence of immunity [12]. Knowing how many women reach delivery unprotected, and where the screening-to-vaccination pathway fails, is needed to target antenatal interventions. This study aimed to determine the prevalence of protective anti-HBs levels and of serological profiles of immunity, infection and susceptibility, and the factors associated with protective anti-HBs levels, among women admitted for delivery at Kawempe National Referral Hospital (KNRH).

## Materials and methods

### Study design and setting

We conducted a hospital-based cross-sectional study. Participants were enrolled between May and June 2024, after administrative clearance had been granted by the hospital. The study is reported according to the Strengthening the Reporting of Observational Studies in Epidemiology (STROBE) guideline [21] (S1 Checklist). KNRH is a public, urban, specialised national referral and teaching hospital in Kampala, Uganda. Its maternity services include antenatal care, delivery, postnatal care, routine immunisation and child growth monitoring. About 2,100 women give birth at the hospital each month, of whom about 25% are referred from other facilities or self-referred.

### Participants and sampling

Pregnant women admitted for delivery at KNRH on data collection days were eligible if they gave written informed consent, irrespective of their hepatitis B vaccination history. Women receiving treatment for HBV infection and women in active labour were excluded. All participants were aged 18 years or older. Participants were selected by systematic random sampling. The sampling interval was calculated by dividing the expected number of admissions for delivery during the data collection period (about 80 per day) by the required sample size, which gave an interval of 3.8, rounded to 4. Every fourth woman admitted on a data collection day was approached; a woman who declined was replaced by the next woman admitted. Participants’ files were marked to prevent re-enrolment.

### Sample size

For the prevalence of protective anti-HBs levels, Kish’s formula [22], n = Z²p(1 − p)/d², with an assumed non-protective prevalence of 22.9% as used in a previous Ugandan study [20], Z = 1.96 and a precision (d) of 5%, gave a minimum of 272 participants. For the analysis of associated factors, a sample size for comparing two proportions computed in G*Power (two-sided α 0.05, 80% power, expected proportions 0.192 and 0.052) gave 383 participants. The larger requirement was adopted, and the enrolment target was set at 420 to allow for incomplete questionnaires or unusable samples.

### Variables

The primary outcome was a protective anti-HBs level, defined as an anti-HBs concentration of at least 10 mIU/mL (10 IU/L) [5]. Women who were HBsAg-negative were further classified by their combination of anti-HBs and hepatitis B core antibody (anti-HBc) results: vaccine-type immunity (anti-HBc-negative, anti-HBs ≥10 mIU/mL), immunity after past infection (anti-HBc-positive, anti-HBs ≥10 mIU/mL), past infection without protective anti-HBs (anti-HBc-positive, anti-HBs <10 mIU/mL) and susceptible (anti-HBc-negative, anti-HBs <10 mIU/mL). Secondary outcomes were the prevalence of HBsAg, HBeAg, hepatitis B e antibody (anti-HBe) and anti-HBc.

The following variables from the questionnaire (S1 File) were analysed: age (recorded in the categories 18–30, 31–40, 41–50 and 51 years or older), education, marital status, residence, previous caesarean delivery, attendance at all scheduled antenatal visits, alcohol use, smoking, self-reported chronic medical condition (including HIV, cancer, sickle cell disease or other chronic disease), previous blood transfusion, previous surgery, having cared for a person with hepatitis B, recreational drug use, multiple sexual partners, sharing of sharp objects, previous hepatitis B testing and its result, and self-reported past hepatitis B. Follow-up items asked only of subgroups (number of antenatal visits among women who had missed visits, reasons for not being vaccinated during the current pregnancy, injection drug use, safer sex practice and initiation of hepatitis B treatment) and two items on perceived availability and quality of antenatal hepatitis B services were not analysed for this paper. Hepatitis B vaccination status was derived from two questions and classified as never vaccinated, vaccinated before the current pregnancy only, or vaccinated during the current pregnancy. Among vaccinated women, completion of three doses, the schedule used and reasons for non-completion were recorded. Vaccination history was self-reported and checked against hospital records where these were available.

### Data collection

A structured questionnaire was pretested among 10 pregnant women at Kajjansi Health Centre IV, Wakiso District, and revised before use. It was administered in face-to-face interviews by the principal investigator and two research assistants (a laboratory technologist and a nurse), who were trained on the tool, quality control and sample handling. The principal investigator reviewed all questionnaires daily for completeness and consistency.

### Laboratory procedures

#### Sample collection and rapid serological screening

From each participant, 4 mL of venous blood was drawn into an ethylenediaminetetraacetic acid (EDTA) tube and centrifuged at 3,000 rpm for 10 minutes, and the plasma was separated into cryovials. Plasma was screened on site with a rapid lateral-flow (immunochromatographic) five-marker hepatitis B test cassette (Dewei Medical Equipment Co. Ltd, China) that qualitatively detects HBsAg, anti-HBs, HBeAg, anti-HBe and anti-HBc. Tests were read visually according to the manufacturer’s instructions and were valid only if the control line appeared. The manufacturer reports a sensitivity of 98.7% and a specificity of 99.1% for HBsAg; performance data for the other markers were not available, and no confirmatory enzyme immunoassay, HBV DNA or alanine aminotransferase testing was performed.

#### Quantitative anti-HBs measurement

The remaining plasma was stored at −80 °C at the Department of Pharmacology and Therapeutics, Makerere University, and transported in cold boxes to the Nakasero Blood Bank laboratory, where anti-HBs was quantified with the Elecsys Anti-HBs assay on an Elecsys 2010 analyser (Roche Diagnostics GmbH, Mannheim, Germany). This is a double-antigen sandwich electrochemiluminescence immunoassay with a measuring range of 2–1,000 IU/L; results of 10 IU/L (equivalent to 10 mIU/mL) or more were classified as protective. The analyser was calibrated and quality control samples were run before each batch, and 50 µL of plasma per sample was analysed according to the manufacturer’s instructions.

### Statistical analysis

#### Descriptive analysis

Data were double-entered in EpiData version 3.1 and analysed in Python version 3.13 with NumPy version 2.5 and SciPy version 1.18. Categorical variables were summarised as frequencies and percentages with numerators and denominators, and prevalences were reported with Wilson 95% confidence intervals (CI). Anti-HBs concentrations were truncated at the limits of the assay measuring range and were not normally distributed, so they were summarised as median and interquartile range (IQR). Characteristics were compared between women with and without protective anti-HBs levels, and between HBsAg-positive and HBsAg-negative women, with Pearson’s chi-square test, or Fisher’s exact test when any expected cell count was below 5 (Monte Carlo estimate with 100,000 samples for tables larger than 2 × 2).

#### Regression analysis

Because the outcome was common, associations with protective anti-HBs levels were estimated as prevalence ratios (PR) with 95% CI using modified Poisson regression, that is, a generalised linear model with a Poisson distribution, log link and robust (sandwich) variance estimator [23]. In the main model (all women), vaccination status, age, smoking and chronic medical condition were included a priori as potential determinants of vaccine response [18], together with all other variables with p < 0.20 in bivariable analysis. Previous hepatitis B testing was not entered because, in the national programme, vaccination follows testing, so testing lies on the pathway to vaccination; serological markers were not entered because they partly define immune status. Because schedule and completion apply only to vaccinated women, these variables were analysed in a separate model restricted to vaccinated women, adjusted for timing of vaccination, age group, smoking and chronic medical condition. As a sensitivity analysis, the main model was repeated among HBsAg-negative women. Analyses were complete-case, and the number of observations in each model is reported. Collinearity among covariates in the main model was assessed with variance inflation factors (VIF), all of which were below 1.7 (S2 File). No interaction or subgroup analyses other than those described were performed. Statistical significance was set at a two-sided p < 0.05. Full regression output is provided in S2 File, and the de-identified dataset with its codebook in S1 Data.

### Ethics statement

The study protocol was approved by the Makerere University School of Biomedical Sciences Research and Ethics Committee (SBS-REC) at its meeting of 16 November 2023 (reference SBS-2023-452; approval valid from 12 February 2024 to 12 February 2025). MO, who chairs the SBS-REC, did not take part in the committee’s review of this protocol. Administrative clearance to conduct the study at KNRH was granted by the Executive Director of the hospital on 17 May 2024 (reference ED.01/04/024). All data were collected after this clearance and within the period of ethics approval. All participants provided written informed consent before enrolment. Under the approved protocol, women found to be infected were to be referred to a physician for further management; vaccination was not provided as part of the study. Personal identifiers were replaced with unique study codes, and the data were stored on password-protected computers accessible only to the study team. The study was conducted in accordance with the Declaration of Helsinki.

## Results

### Participants and vaccination history

A total of 418 women were enrolled, against a target of 420; refusals were not recorded in the study database. Most were aged 18–40 years (338/418, 80.9%), 187 (44.7%) had tertiary education and 250 (59.8%) lived in urban areas (Table 1). Data were missing for multiple sexual partners (26 women), recreational drug use (17), sharing of sharp objects (6) and antenatal attendance (3).

**Table 1.** Characteristics of participants by hepatitis B surface antibody (anti-HBs) status, Kawempe National Referral Hospital, 2024 (N = 418).

| **Characteristic** | **Category** | **Total, n (%)** | **Anti-HBs ≥10 mIU/mL, n (%)** | **Anti-HBs <10 mIU/mL, n (%)** | **χ² (df)** | **p-value** |
|---|---|---|---|---|---|---|
| Age (years) | 18–30 | 160 (38.3) | 120 (75.0) | 40 (25.0) | – | 0.87<sup>a</sup> |
|  | 31–40 | 178 (42.6) | 140 (78.7) | 38 (21.3) |  |  |
|  | 41–50 | 69 (16.5) | 53 (76.8) | 16 (23.2) |  |  |
|  | ≥51 | 11 (2.6) | 9 (81.8) | 2 (18.2) |  |  |
| Education | No formal education | 100 (23.9) | 85 (85.0) | 15 (15.0) | – | 0.07<sup>a</sup> |
|  | Primary | 18 (4.3) | 16 (88.9) | 2 (11.1) |  |  |
|  | Secondary | 113 (27.0) | 85 (75.2) | 28 (24.8) |  |  |
|  | Tertiary | 187 (44.7) | 136 (72.7) | 51 (27.3) |  |  |
| Marital status | Single | 108 (25.8) | 83 (76.9) | 25 (23.1) | 1.22 (3) | 0.75 |
|  | Married | 157 (37.6) | 124 (79.0) | 33 (21.0) |  |  |
|  | Divorced | 97 (23.2) | 71 (73.2) | 26 (26.8) |  |  |
|  | Widowed | 56 (13.4) | 44 (78.6) | 12 (21.4) |  |  |
| Residence | Urban | 250 (59.8) | 190 (76.0) | 60 (24.0) | 0.38 (1) | 0.54 |
|  | Rural | 168 (40.2) | 132 (78.6) | 36 (21.4) |  |  |
| Previous caesarean delivery | Yes | 108 (25.8) | 97 (89.8) | 11 (10.2) | 13.45 (1) | <0.001 |
|  | No | 310 (74.2) | 225 (72.6) | 85 (27.4) |  |  |
| Attended all antenatal visits (missing 3) | Yes | 255 (61.0) | 212 (83.1) | 43 (16.9) | 14.62 (1) | <0.001 |
|  | No | 160 (38.3) | 107 (66.9) | 53 (33.1) |  |  |
| Alcohol use | Yes | 101 (24.2) | 78 (77.2) | 23 (22.8) | 0.00 (1) | 0.96 |
|  | No | 317 (75.8) | 244 (77.0) | 73 (23.0) |  |  |
| Smoking | Yes | 60 (14.4) | 50 (83.3) | 10 (16.7) | 1.57 (1) | 0.21 |
|  | No | 358 (85.6) | 272 (76.0) | 86 (24.0) |  |  |
| Chronic medical condition | Yes | 73 (17.5) | 51 (69.9) | 22 (30.1) | 2.57 (1) | 0.11 |
|  | No | 345 (82.5) | 271 (78.6) | 74 (21.4) |  |  |
| Previous blood transfusion | Yes | 102 (24.4) | 88 (86.3) | 14 (13.7) | 6.51 (1) | 0.01 |
|  | No | 316 (75.6) | 234 (74.1) | 82 (25.9) |  |  |
| Previous surgery | Yes | 100 (23.9) | 91 (91.0) | 9 (9.0) | 14.49 (1) | <0.001 |
|  | No | 318 (76.1) | 231 (72.6) | 87 (27.4) |  |  |
| Cared for a person with hepatitis B | Yes | 114 (27.3) | 93 (81.6) | 21 (18.4) | 1.83 (1) | 0.18 |
|  | No | 304 (72.7) | 229 (75.3) | 75 (24.7) |  |  |
| Recreational drug use (missing 17) | Yes | 37 (8.9) | 29 (78.4) | 8 (21.6) | 0.03 (1) | 0.87 |
|  | No | 364 (87.1) | 281 (77.2) | 83 (22.8) |  |  |
| Multiple sexual partners (missing 26) | Yes | 94 (22.5) | 65 (69.1) | 29 (30.9) | 4.05 (1) | 0.04 |
|  | No | 298 (71.3) | 236 (79.2) | 62 (20.8) |  |  |
| Shared sharp objects (missing 6) | Yes | 77 (18.4) | 55 (71.4) | 22 (28.6) | 1.62 (1) | 0.20 |
|  | No | 335 (80.1) | 262 (78.2) | 73 (21.8) |  |  |
| Previously tested for hepatitis B | Yes | 349 (83.5) | 300 (86.0) | 49 (14.0) | 95.22 (1) | <0.001 |
|  | No | 69 (16.5) | 22 (31.9) | 47 (68.1) |  |  |
| Self-reported past hepatitis B | Yes | 52 (12.4) | 41 (78.8) | 11 (21.2) | 0.11 (1) | 0.74 |
|  | No | 366 (87.6) | 281 (76.8) | 85 (23.2) |  |  |
| Hepatitis B vaccination | Never | 154 (36.8) | 78 (50.6) | 76 (49.4) | 100.84 (2) | <0.001 |
|  | Before current pregnancy only | 61 (14.6) | 50 (82.0) | 11 (18.0) |  |  |
|  | During current pregnancy | 203 (48.6) | 194 (95.6) | 9 (4.4) |  |  |
| HBsAg (rapid test) | Positive | 21 (5.0) | 15 (71.4) | 6 (28.6) | – | 0.59<sup>a</sup> |
|  | Negative | 397 (95.0) | 307 (77.3) | 90 (22.7) |  |  |
| Anti-HBc (rapid test) | Positive | 39 (9.3) | 28 (71.8) | 11 (28.2) | 0.67 (1) | 0.41 |
|  | Negative | 379 (90.7) | 294 (77.6) | 85 (22.4) |  |  |

Total percentages are column percentages; percentages in the anti-HBs columns are row percentages (proportion of each category with or without protective levels). χ² (df), Pearson chi-square statistic with degrees of freedom; p-values are from Pearson's chi-square test, or <sup>a</sup> Fisher's exact test (Monte Carlo estimate for tables larger than 2 × 2), for which no test statistic is shown (–). Chronic medical condition includes HIV, cancer, sickle cell disease and other chronic diseases (self-reported). HBsAg and anti-HBc were measured with a rapid test. anti-HBc, hepatitis B core antibody; HBsAg, hepatitis B surface antigen.

Overall, 349 women (83.5%) reported a previous hepatitis B test; 300 reported a negative result and 49 a positive result. In total, 264 women (63.2%; 95% CI 58.4–67.6) reported hepatitis B vaccination: 203 (48.6%) during the current pregnancy and 61 (14.6%) before the current pregnancy only. Of the 154 never-vaccinated women, 50 had reported a previous negative test, 57 had never been tested and 47 had reported a previous positive test. Thus, 50 of the 300 women (16.7%) who had tested negative had not been vaccinated (Fig 1).

**Fig 1.**
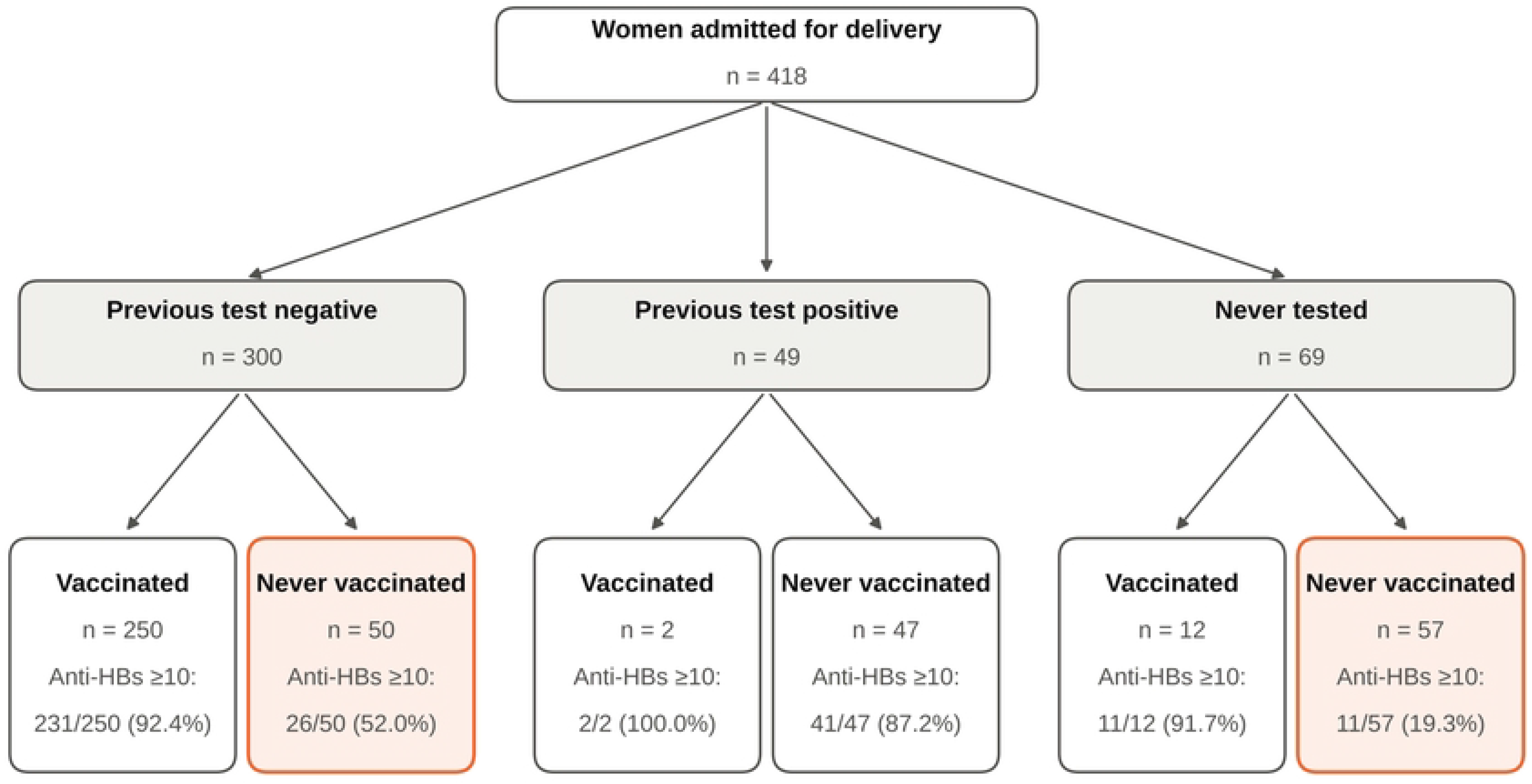
Hepatitis B testing, vaccination and protective antibody levels among women admitted for delivery. Women (N = 418) are grouped by their self-reported previous hepatitis B test result and by vaccination history (vaccinated before or during the current pregnancy, or never vaccinated). Each lower box shows the number and proportion of women with protective hepatitis B surface antibody (anti-HBs) levels (≥10 mIU/mL). Highlighted boxes show never-vaccinated women who had tested negative, for whom a negative test was not followed by vaccination, and never-vaccinated women who had never been tested.

Of the 264 vaccinated women, 182 (68.9%) reported completing three doses, 68 (25.8%) had not completed the series and 14 (5.3%) did not report completion. Among the 68 women with an incomplete series, the reasons were not being told to return for the next dose (30, 44.1%), forgetting (23, 33.8%), vaccine stock-out (9, 13.2%), other reasons (4, 5.9%) and not stated (2, 2.9%). Of the 199 vaccinated women who reported a schedule, 183 (92.0%) followed the 0, 1 and 6 month schedule.

### Anti-HBs levels and serological profiles

The median anti-HBs concentration was 328.4 mIU/mL (IQR 20.4–561.7). Overall, 322 women (77.0%; 95% CI 72.8–80.8) had protective levels. Protection was found in 78 of 154 never-vaccinated women (50.6%), 50 of 61 women vaccinated before the current pregnancy only (82.0%) and 194 of 203 women vaccinated during the current pregnancy (95.6%) (Table 1 and Fig 2). Among vaccinated women, 170 of 182 (93.4%) who had completed three doses and 60 of 68 (88.2%) who had not had protective levels.

**Fig 2.**
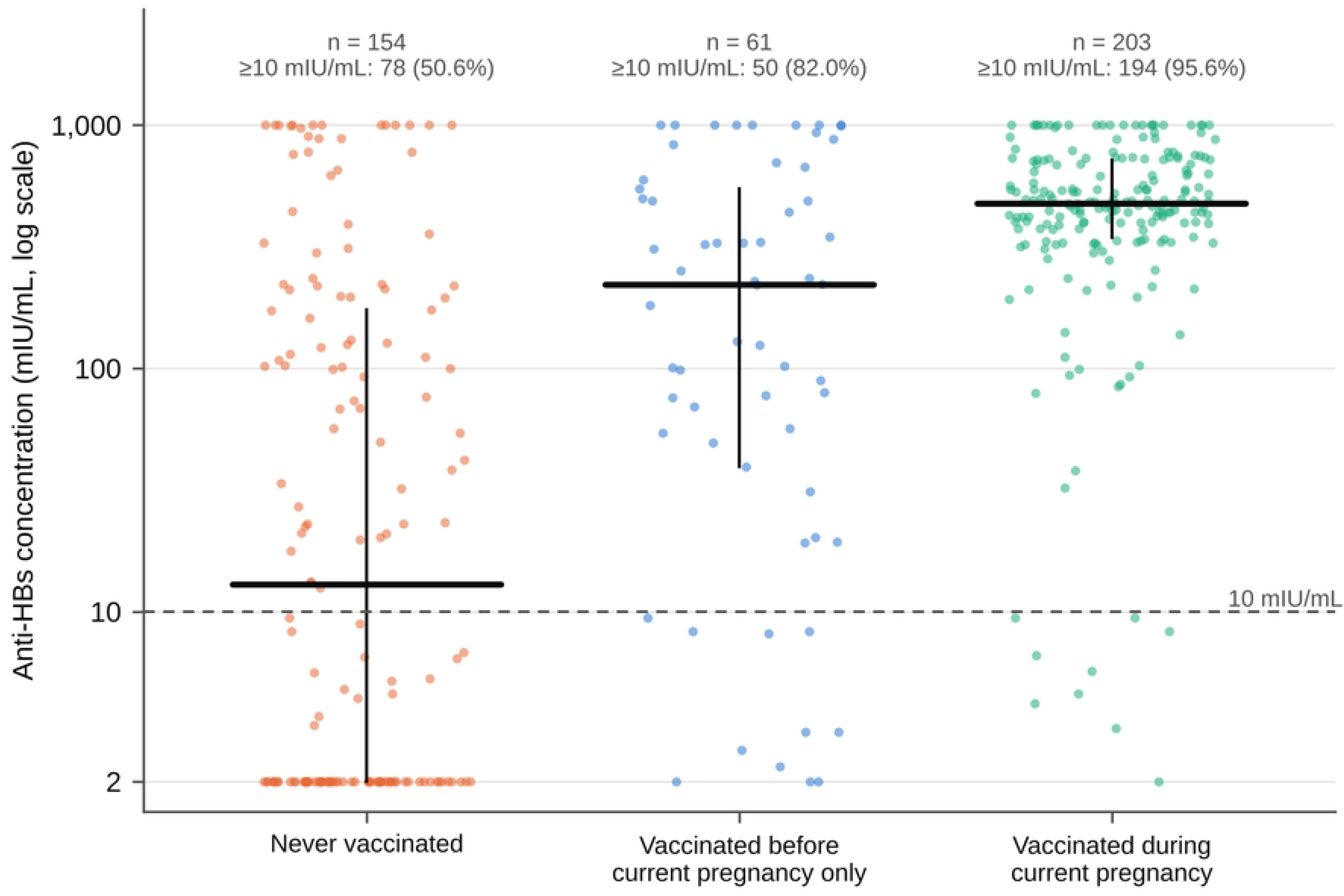
Anti-HBs concentrations by hepatitis B vaccination status among women admitted for delivery. Each dot represents one woman (N = 418); horizontal bars show medians and vertical lines interquartile ranges. Concentrations are plotted on a log scale and truncated at the limits of the assay measuring range (2–1,000 mIU/mL). The dashed line marks the protective threshold of 10 mIU/mL. Median concentrations were 12.9 mIU/mL among never-vaccinated women, 219.7 mIU/mL among women vaccinated before the current pregnancy only and 474.8 mIU/mL among women vaccinated during the current pregnancy. anti-HBs, hepatitis B surface antibody.

The rapid test detected HBsAg in 21 women (5.0%; 95% CI 3.3–7.6), HBeAg in 19 (4.5%; 95% CI 2.9–7.0), anti-HBe in 50 (12.0%; 95% CI 9.2–15.4) and anti-HBc in 39 (9.3%; 95% CI 6.9–12.5). Several results were biologically discordant. Fifteen of the 19 HBeAg-positive women were HBsAg-negative, 30 of the 50 anti-HBe-positive women were anti-HBc-negative, and 15 of the 21 HBsAg-positive women had anti-HBs of at least 10 mIU/mL. In addition, only 2 of the 49 women who reported a previous positive test were HBsAg-positive in this study, whereas 14 of the 21 HBsAg-positive women had reported a previous negative test.

Among the 397 HBsAg-negative women, 287 (72.3%) had vaccine-type immunity, 20 (5.0%) had immunity after past infection, 8 (2.0%) had anti-HBc without protective anti-HBs, and 82 (20.7%; 95% CI 17.0–24.9) were susceptible (Table 2). The 82 susceptible women made up 19.6% of all participants; 65 of them (79.3%) had never been vaccinated. Among never-vaccinated women, 74 of 154 (48.1%) had a vaccine-type immunity profile.

**Table 2.** Serological profiles by hepatitis B vaccination status among women admitted for delivery, Kawempe National Referral Hospital, 2024 (N = 418).

| **Serological profile** | **Interpretation** | **All (n = 418)** | **Never vaccinated (n = 154)** | **Vaccinated before pregnancy only (n = 61)** | **Vaccinated during pregnancy (n = 203)** |
|---|---|---|---|---|---|
| HBsAg−, anti-HBc−, anti-HBs ≥10 | Vaccine-type immunity | 287 (68.7) | 74 (48.1) | 41 (67.2) | 172 (84.7) |
| HBsAg−, anti-HBc+, anti-HBs ≥10 | Immunity after past infection | 20 (4.8) | 2 (1.3) | 5 (8.2) | 13 (6.4) |
| HBsAg−, anti-HBc+, anti-HBs <10 | Past infection, non-protective anti-HBs | 8 (1.9) | 6 (3.9) | 0 (0.0) | 2 (1.0) |
| HBsAg−, anti-HBc−, anti-HBs <10 | Susceptible | 82 (19.6) | 65 (42.2) | 10 (16.4) | 7 (3.4) |
| HBsAg positive | Current infection | 21 (5.0) | 7 (4.5) | 5 (8.2) | 9 (4.4) |

Values are n (column %). anti-HBs was measured quantitatively (≥10 mIU/mL considered protective); HBsAg and anti-HBc were measured with a rapid test. Susceptible: no serological evidence of immunity or infection. A vaccinated woman with anti-HBs <10 mIU/mL may retain immune memory. anti-HBc, hepatitis B core antibody; anti-HBs, hepatitis B surface antibody; HBsAg, hepatitis B surface antigen.

### Factors associated with protective anti-HBs levels

In the main model (389 women with complete data), vaccination before the current pregnancy only (adjusted PR [aPR] 1.54; 95% CI 1.24–1.89; p < 0.001) and vaccination during the current pregnancy (aPR 1.86; 95% CI 1.57–2.20; p < 0.001) were associated with protective levels, compared with no vaccination (Table 3). Previous caesarean delivery, previous surgery, previous blood transfusion, attendance at all antenatal visits and education were associated with protection in bivariable analysis, but none remained significant after adjustment. Age, smoking and chronic medical condition were not associated with protection. The results were unchanged when the analysis was restricted to the 370 HBsAg-negative women with complete data (vaccination before the current pregnancy only: aPR 1.53, 95% CI 1.23–1.90; during the current pregnancy: aPR 1.86, 95% CI 1.56–2.21; S2 File).

**Table 3.**
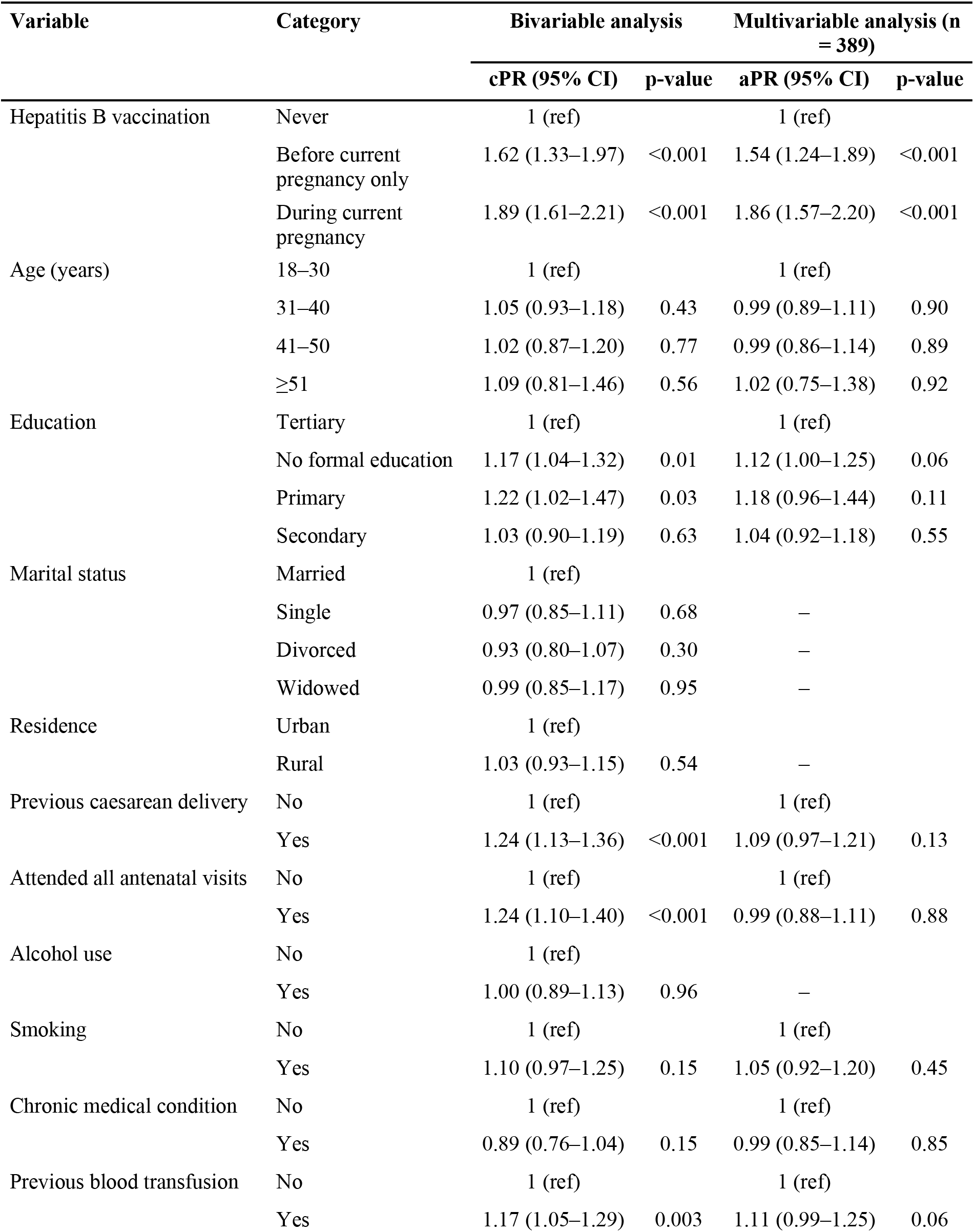
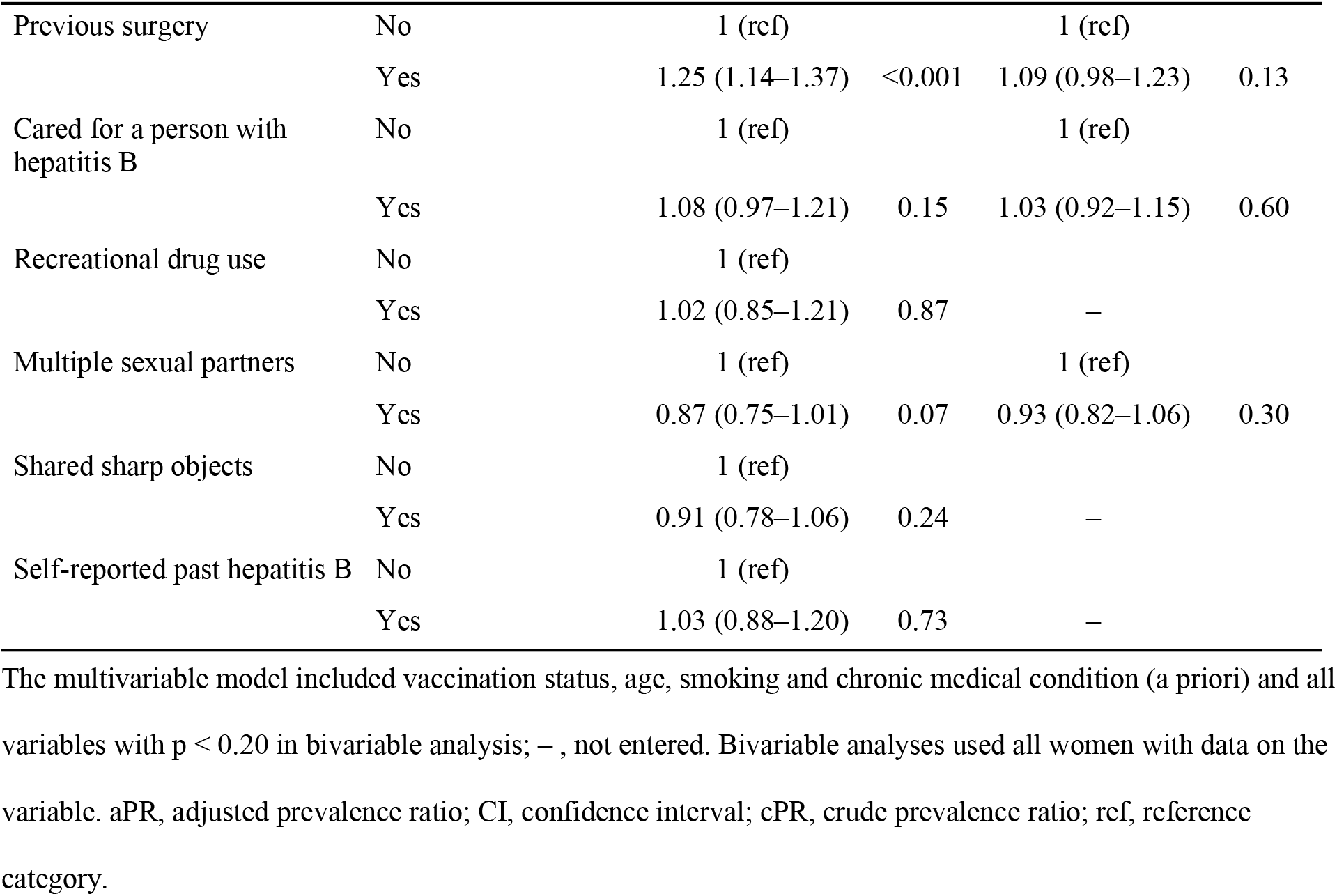
Factors associated with protective hepatitis B surface antibody levels (≥10 mIU/mL) among women admitted for delivery: modified Poisson regression, all women.

Among vaccinated women (250 with complete data), vaccination during the current pregnancy was associated with a higher prevalence of protection than vaccination before it only (aPR 1.17; 95% CI 1.04–1.33; p = 0.01), whereas completion of three doses was not significantly associated (aPR 1.07; 95% CI 0.98–1.18; p = 0.13) (Table 4). The schedule could not be modelled, because all 16 women on schedules other than 0, 1 and 6 months had protective levels.

**Table 4.** Factors associated with protective hepatitis B surface antibody levels among vaccinated women (n = 264): modified Poisson regression.

| Variable | Category | Protective, n/N (%) | cPR (95% CI) | p-value | aPR (95% CI), n = 250 | p-value |
|---|---|---|---|---|---|---|
| Timing of vaccination | Before current pregnancy only | 50/61 (82.0) | 1 (ref) |  | 1 (ref) |  |
|  | During current pregnancy | 194/203 (95.6) | 1.17 (1.03–1.32) | 0.01 | 1.17 (1.04–1.33) | 0.01 |
| Completed three doses (missing 14) | No | 60/68 (88.2) | 1 (ref) |  | 1 (ref) |  |
|  | Yes | 170/182 (93.4) | 1.06 (0.96–1.16) | 0.24 | 1.07 (0.98–1.18) | 0.13 |
| Age (years) | 18–30 |  | 1 (ref) |  | 1 (ref) |  |
|  | 31–40 |  | 1.01 (0.94–1.10) | 0.75 | 1.02 (0.94–1.11) | 0.64 |
|  | ≥41 |  | 1.01 (0.92–1.11) | 0.85 | 1.04 (0.94–1.15) | 0.42 |
| Smoking | No |  | 1 (ref) |  | 1 (ref) |  |
|  | Yes |  | 1.00 (0.91–1.10) | 0.94 | 0.99 (0.90–1.10) | 0.92 |
| Chronic medical condition | No |  | 1 (ref) |  | 1 (ref) |  |
|  | Yes |  | 0.96 (0.85–1.08) | 0.49 | 0.96 (0.86–1.08) | 0.50 |

The multivariable model included all variables shown. Vaccination schedule was not modelled because all 16 women on schedules other than 0, 1 and 6 months had protective levels. aPR, adjusted prevalence ratio; CI, confidence interval; cPR, crude prevalence ratio; ref, reference category.

HBsAg positivity did not differ by vaccination status or most other characteristics, but it was more frequent among women who reported sharing sharp objects (11/77, 14.3%) than among those who did not (10/335, 3.0%; p < 0.001) (S1 Table).

## Discussion

In this study of women admitted for delivery at a national referral hospital in Uganda, 77.0% had protective anti-HBs levels, but one in five (19.6%) had no serological evidence of either immunity or infection and was therefore susceptible to HBV. Four in five of these susceptible women had never been vaccinated. Vaccination was strongly associated with protection, and almost all women vaccinated during the current pregnancy were protected. No socio-demographic or behavioural characteristic was independently associated with protection.

### Why women reached delivery unprotected

Our data show where the pathway from testing to protection breaks down. The largest gap was failure to vaccinate. One in six women who reported a negative hepatitis B test had never been vaccinated, and 57 unvaccinated women had never been tested at all, despite the availability of free adult screening and vaccination since 2015 [14]. Women who had been tested were far more likely to be protected than those who had not (86.0% vs 31.9%; Table 1), consistent with testing being the usual route to vaccination. Associations of protection with previous surgery, caesarean delivery, blood transfusion and full antenatal attendance in bivariable analysis disappeared after adjustment for vaccination. This suggests that contact with health services matters mainly because it creates opportunities for testing and vaccination. Offering the first vaccine dose at the antenatal visit at which a negative result is returned would address this gap directly.

The second gap was an incomplete vaccine series. A quarter of vaccinated women had not completed three doses, and the reasons they gave were mostly health-system failures rather than refusal: they were not told to return, they forgot, or the vaccine was out of stock. Similar barriers, together with vaccine hesitancy, have been described among pregnant women in Kampala, in other Ugandan populations [16,24] and across sub-Saharan Africa [15]. Most women followed the 0, 1 and 6 month schedule, which cannot be completed before delivery if it is started after the first trimester. Schedules with shorter intervals between doses are acceptable for adults [5,17]. Protection was more frequent among women vaccinated during the current pregnancy than among those vaccinated only before it. This is consistent with a recent dose producing higher antibody concentrations, which then decline over time [19,25]. A concentration below 10 mIU/mL in a previously vaccinated woman does not necessarily mean loss of protection, because people who responded to vaccination usually retain immune memory [5,19]. The 17 vaccinated women classified as susceptible may therefore overstate true susceptibility. Completion of three doses was not significantly associated with protection, possibly because of inaccurate recall of the number of doses. Hepatitis B vaccination during pregnancy is safe [26].

### Protection among women who reported no vaccination

The proportion of women with protective levels (77.0%) exceeded the proportion who reported vaccination (63.2%) because half of the never-vaccinated women had protective levels. Almost all of these women had a vaccine-type profile (anti-HBc-negative), so natural infection explains little of this immunity, although rapid anti-HBc testing may miss some past infections. The most likely explanations are vaccination that women did not report or remember, including infant vaccination in the youngest women through the pentavalent vaccine [10], and misclassification of vaccination status. This finding indicates that self-reported vaccination history is an unreliable guide to protection in this population.

### Hepatitis B infection markers

The rapid test detected HBsAg in 5.0% of women, which is close to the pooled estimate of 4.9% for pregnant women in East Africa [7]. It is higher than the 3.0% reported among women nationally [9] and the prevalence found among pregnant women in south-western Uganda [11,12], but lower than the 11.8% reported in northern Uganda [10]. However, this estimate must be interpreted with caution. Most HBsAg-positive women also had protective anti-HBs, which is uncommon in true chronic infection. Most HBeAg-positive women were HBsAg-negative, and anti-HBe was often found without anti-HBc; both patterns are biologically implausible. Rapid HBsAg results also agreed poorly with women’s reports of previous tests. Together, these findings suggest that the rapid multi-marker test produced false-positive results, particularly for HBeAg and anti-HBe. Because no confirmatory immunoassay or HBV DNA testing was performed, we cannot estimate the true prevalence of infection or the risk of transmission. These results support WHO’s recommendation that reactive rapid tests be confirmed before women are classified as infected and assessed for antiviral prophylaxis [6]. The association between sharing sharp objects and HBsAg positivity is consistent with blood-borne transmission but is based on few cases.

### Implications

This study did not evaluate interventions, so the following implications are based on the gaps we observed. The main avoidable gap was that a negative test was not consistently followed by vaccination. Linking every negative antenatal test to a first vaccine dose at the same visit, with reminders and a reliable vaccine supply, would directly address it. Now that the hepatitis B birth dose is part of Uganda’s routine schedule [13], newborns can be protected regardless of their mothers’ immunity; antenatal testing therefore remains essential mainly to identify infected mothers, using confirmed results, so that eligible mothers receive antiviral prophylaxis [6].

### Strengths and limitations

Strengths of this study include systematic random sampling, quantitative measurement of anti-HBs with an automated immunoassay, cross-classification of anti-HBs with anti-HBc to distinguish vaccine-type from natural immunity, and reporting of all exposure and risk-factor variables collected. The study also has limitations. First, its cross-sectional design does not allow causal inference, and it was conducted at a single national referral hospital, so the findings may not be generalisable to women delivering at lower-level facilities. Second, vaccination history and exposures were self-reported. Vaccination cards were not systematically available, and the high prevalence of protection among never-vaccinated women indicates substantial misclassification. Third, HBsAg, HBeAg, anti-HBe and anti-HBc were detected with a rapid test whose performance for markers other than HBsAg has not been validated, without confirmatory immunoassay, HBV DNA or alanine aminotransferase testing, and the discordant patterns we observed indicate false-positive results. Fourth, HIV status was recorded only within a composite question on chronic illness, and body mass index, parity, diabetes and time since the last vaccine dose were not available, so residual confounding is possible. Fifth, age was recorded in broad categories, and 11 women reported being 51 years or older, which may reflect misreporting. Sixth, a single anti-HBs measurement cannot distinguish susceptible women from vaccinated women with waning antibody but intact immune memory. Seventh, we did not record whether HBsAg-positive women were linked to care or whether their infants received prophylaxis, or whether susceptible women were later vaccinated. Finally, refusals were not recorded, so participants could not be compared with women who declined.

## Conclusions

Most women delivering at KNRH had protective anti-HBs levels, but one in five reached delivery with no serological evidence of immunity or infection, and most of these women had never been vaccinated. The main avoidable gaps were that a negative test was not consistently followed by vaccination and that started vaccine series were not completed, largely for health-system reasons. Linking every negative antenatal hepatitis B test to vaccination, and confirming reactive rapid tests before women are classified as infected, could strengthen efforts to eliminate mother-to-child transmission of HBV in Uganda.

## Data Availability

All relevant data are within the manuscript and its Supporting Information files. The de-identified participant-level dataset and codebook are provided as S1 Data..

## Acknowledgments

We thank the women who took part in this study. We also thank the management and staff of Kawempe National Referral Hospital, in particular the research assistants (a laboratory technologist and a nurse) who supported participant recruitment, interviews and sample collection, and the staff of the Nakasero Blood Bank laboratory who performed the anti-HBs assays. We are grateful to the Department of Pharmacology and Therapeutics, School of Biomedical Sciences, Makerere University, for its support.

## Supporting information

**S1 Checklist. STROBE checklist for cross-sectional studies.** Completed STROBE checklist indicating where each item is reported in the manuscript.

**S1 File. Study questionnaire (English).** Interviewer-administered structured questionnaire used for data collection, with the corresponding dataset variable names.

**S1 Table. Hepatitis B surface antigen positivity by participant characteristics.** Prevalence of hepatitis B surface antigen (rapid test) by socio-demographic, clinical, behavioural and vaccination characteristics, with p-values.

**S2 File. Full output of the modified Poisson regression models.** Coefficients, robust standard errors, prevalence ratios with 95% confidence intervals and p-values for the bivariable, main multivariable, sensitivity (HBsAg-negative women) and vaccinated-women models.

**S1 Data. De-identified dataset with codebook.** De-identified participant-level dataset (N = 418) with a codebook describing every variable and its coding (Excel workbook).

## References

1. World Health Organization. Global hepatitis report 2024: action for access in low- and middle-income countries. Geneva: World Health Organization; 2024.

2. Polaris Observatory Collaborators. Global prevalence, cascade of care, and prophylaxis coverage of hepatitis B in 2022: a modelling study. Lancet Gastroenterol Hepatol. 2023;8(10):879–907. doi: 10.1016/S2468-1253(23)00197-8

3. Veronese P, Dodi I, Esposito S, Indolfi G. Prevention of vertical transmission of hepatitis B virus infection. World J Gastroenterol. 2021;27(26):4182–93. doi: 10.3748/wjg.v27.i26.4182

4. Joshi SS, Coffin CS. Hepatitis B and pregnancy: virologic and immunologic characteristics. Hepatol Commun. 2020;4(2):157–71. doi: 10.1002/hep4.1460

5. World Health Organization. Hepatitis B vaccines: WHO position paper – July 2017. Wkly Epidemiol Rec. 2017;92(27):369–92.

6. World Health Organization. Guidelines for the prevention, diagnosis, care and treatment for people with chronic hepatitis B infection. Geneva: World Health Organization; 2024.

7. Larebo YM, Anshebo AA, Abdo RA, Behera SK, Gopalan N. Prevalence of hepatitis B virus infection among pregnant women in Africa: a systematic review and meta-analysis. PLoS One. 2024;19(7):e0305838. doi: 10.1371/journal.pone.0305838

8. Bwogi J, Braka F, Makumbi I, Mishra V, Bakamutumaho B, Nanyunja M, et al. Hepatitis B infection is highly endemic in Uganda: findings from a national serosurvey. Afr Health Sci. 2009;9(2):98–108.

9. Uganda Ministry of Health. Uganda Population-based HIV Impact Assessment (UPHIA) 2016–2017: summary sheet of preliminary findings. Kampala: Ministry of Health; 2017.

10. Bayo P, Ochola E, Oleo C, Mwaka AD. High prevalence of hepatitis B virus infection among pregnant women attending antenatal care: a cross-sectional study in two hospitals in northern Uganda. BMJ Open. 2014;4(11):e005889. doi: 10.1136/bmjopen-2014-005889

11. Mugabiirwe N, Kalyetsi R, Ayella R, Obote J, Ssedyabane F. Hepatitis B virus infection and HBeAg positivity among pregnant women in South West Uganda. Afr J Lab Med. 2022;11(1):a1784. doi: 10.4102/ajlm.v11i1.1784

12. Kabajulizi I, Bazira J, Atuheire C, Kato C, Kabanda T. Hepatitis B infection and immunity among pregnant women attending antenatal clinics in health centers of Mbarara Municipality, southwestern Uganda. Adv Infect Dis. 2019;9(2):65–79. doi: 10.4236/aid.2019.92006

13. Musenze J. Uganda rolls out the hepatitis B birth dose vaccine, to protect the youngest and most vulnerable [Internet]. Geneva: Gavi, the Vaccine Alliance; 2025 Oct 31 [cited 2026 Nov 3]. Available from: https://www.gavi.org/vaccineswork/uganda-rolls-out-hepatitis-b-birth-dose-vaccine-protect-youngest-and-most

14. World Health Organization. Uganda elevates health for all as the population accesses free hepatitis testing, treatment and vaccines [Internet]. Geneva: World Health Organization; 2023 Jul 28 [cited 2026 Nov 3]. Available from: https://www.who.int/news-room/feature-stories/detail/uganda-elevates-health-for-all-as-the-population-accesses-free-hepatitis-testing--treatment-and-vaccines

15. Boisson A, Goel V, Yotebieng M, Parr JB, Fried B, Thompson P. Implementation approaches for introducing and overcoming barriers to hepatitis B birth-dose vaccine in sub-Saharan Africa. Glob Health Sci Pract. 2022;10(1):e2100277. doi: 10.9745/GHSP-D-21-00277

16. Afolabi IB, Aremu AB, Maidoki LA, Atulomah NO. Predictors of hepatitis B virus infection vaccine hesitancy among pregnant women attending antenatal care at Lubaga Hospital, Kampala, Uganda. Int J Womens Health. 2022;14:1093–104. doi: 10.2147/IJWH.S378000

17. Schillie S, Vellozzi C, Reingold A, Harris A, Haber P, Ward JW, et al. Prevention of hepatitis B virus infection in the United States: recommendations of the Advisory Committee on Immunization Practices. MMWR Recomm Rep. 2018;67(1):1–31. doi: 10.15585/mmwr.rr6701a1

18. Yang S, Tian G, Cui Y, Ding C, Deng M, Yu C, et al. Factors influencing immunologic response to hepatitis B vaccine in adults. Sci Rep. 2016;6:27251. doi: 10.1038/srep27251

19. Bruce MG, Bruden D, Hurlburt D, Zanis C, Thompson G, Rea L, et al. Antibody levels and protection after hepatitis B vaccine: results of a 30-year follow-up study and response to a booster dose. J Infect Dis. 2016;214(1):16–22. doi: 10.1093/infdis/jiv748

20. Ocan M, Acheng F, Otike C, Beinomugisha J, Katete D, Obua C. Antibody levels and protection after hepatitis B vaccine in adult vaccinated healthcare workers in northern Uganda. PLoS One. 2022;17(1):e0262126. doi: 10.1371/journal.pone.0262126

21. von Elm E, Altman DG, Egger M, Pocock SJ, Gøtzsche PC, Vandenbroucke JP. The Strengthening the Reporting of Observational Studies in Epidemiology (STROBE) statement: guidelines for reporting observational studies. PLoS Med. 2007;4(10):e296. doi: 10.1371/journal.pmed.0040296

22. Kish L. Survey sampling. New York: John Wiley and Sons; 1965.

23. Zou G. A modified Poisson regression approach to prospective studies with binary data. Am J Epidemiol. 2004;159(7):702–6. doi: 10.1093/aje/kwh090

24. Ssekamatte T, Isunju JB, Mutyoba JN, Tetui M, Mugambe RK, Nalugya A, et al. Predictors of hepatitis B screening and vaccination status of young psychoactive substance users in informal settlements in Kampala, Uganda. PLoS One. 2022;17(5):e0267953. doi: 10.1371/journal.pone.0267953

25. Zhao YL, Han BH, Zhang XJ, Pan LL, Zhou HS, Gao Z, et al. Immune persistence 17 to 20 years after primary vaccination with recombination hepatitis B vaccine (CHO) and the effect of booster dose vaccination. BMC Infect Dis. 2019;19(1):482. doi: 10.1186/s12879-019-4134-9

26. Groom HC, Irving SA, Koppolu P, Smith N, Vazquez-Benitez G, Kharbanda EO, et al. Uptake and safety of hepatitis B vaccination during pregnancy: a Vaccine Safety Datalink study. Vaccine. 2018;36(41):6111–6. doi: 10.1016/j.vaccine.2018.08.074

